# Dietary Inflammatory Potential and Its Association with Asthma and Lung Function in a Korean Adult Population

**DOI:** 10.64898/2026.03.20.26348946

**Authors:** Da Seul Park, Jongseok Lee, Hansol Lee, Soo Jie Chung, Jin-ah Sim

## Abstract

**Background:** Diets with high inflammatory potential may contribute to asthma and impaired lung function, yet evidence from Asian populations is limited.

**Objective:** We aimed to examine the association between the energy-adjusted Dietary Inflammatory Index (E-DII) and lung function in Korean adults, stratified by asthma status.

**Methods:** Data were analyzed from 12,400 participants in the Korea National Health and Nutrition Examination Survey (2016–2018). The E-DII was calculated from 24-hour dietary recall using 21 validated food parameters. Lung function (FEV₁, FVC) was measured by standardized spirometry, and current asthma was defined as both a physician diagnosis and the presence of current symptoms. Multivariable logistic and linear regression models adjusted for age and sex were applied.

**Results:** The mean age of participants was 49.8 years, and 51.3% were female; 246 (2.0%) reported current asthma. Compared with non-asthmatic individuals, those with asthma were older and more often female. Higher E-DII scores were associated with greater asthma prevalence and reduced lung function, with stronger associations observed among participants with asthma.

**Conclusions:** Higher dietary inflammatory potential was linked to increased asthma risk and lower lung function in Korean adults. These findings suggest that reducing dietary inflammation may benefit respiratory health, although longitudinal studies are needed to confirm temporality and causal direction.

## INTRODUCTION

Asthma is a chronic respiratory condition characterized by airway inflammation, respiratory symptoms, and expiratory airflow limitation. It remains a major global health concern, affecting an estimated 262 million individuals and causing over 455,000 deaths annually as of 2019 [1]. The prevalence of asthma is particularly high in developed countries, ranging from 15% to 20%, and is steadily increasing in developing regions [2]. This global burden may be partly attributable to environmental and lifestyle changes, among which dietary transition toward a westernized pattern is particularly important. Such diets, typically high in saturated fats and processed foods, have been associated with airway inflammation and altered immune responses that may promote allergic disease development [3,4].

A meta-analysis has shown that higher intake of vitamins C, E, and D, as well as fruit consumption and adherence to the Mediterranean diet, are inversely associated with asthma and wheezing [5]. These protective associations may be explained in part by their ability to modulate airway inflammation, as asthma is characterized by chronic inflammation involving eosinophils, mast cells, and T-helper 2 (Th2) lymphocytes, which produce cytokines such as IL-4, IL-5, and IL-13 that drive disease development [6]. However, the effects of individual nutrients are often modest, and synergistic interactions between dietary components are frequently overlooked, highlighting the importance of evaluating overall dietary patterns in asthma research [7].

The Dietary Inflammatory Index (DII) is a literature-derived scoring system that estimates the inflammatory potential of an individual’s diet. Higher DII scores reflect more pro-inflammatory diets [8]. A revised version, the energy-adjusted DII (E-DII), addresses differences in total energy intake by standardizing food component intake to 1000 kcal, improving its applicability in population studies [9]. Prior studies have suggested that pro-inflammatory diets may impair lung function [10]. Considering the central role of inflammation in asthma, it is reasonable to infer that such diets may also contribute to the development or exacerbation of asthma.

Despite this, large-scale real-world investigations evaluating the relationship between E-DII scores, asthma prevalence, and lung function remain scarce. This study seeks to fill this gap by examining the association between E-DII scores and asthma diagnosis and lung function using nationally representative Korean health data. Through this analysis, we aim to clarify the role of pro-inflammatory dietary patterns in respiratory health and provide evidence for potential dietary interventions in asthma management.

## MATERIALS AND METHODS

### Study population

This study analyzed data from 32,379 individuals who participated in the Korea National Health and Nutrition Examination Survey (KNHANES) from 2016 to 2018. The recruitment period for the participants of the KNHANES (2016–2018) used in this study spanned from January 2016 to December 2018. KNHANES is a nationally representative, cross-sectional survey designed to assess the health and nutritional status of the Korean population [11]. All participants provided written informed consent prior to their participation in the survey, and the consent process was conducted by trained staff from the Korea Disease Control and Prevention Agency (KDCA). The survey protocols and procedures of the KNHANES were approved by the Institutional Review Board of the KDCA (approval no. 2018-01-03-P-A). Participants were excluded if they had missing data on dietary intake based on the 24-hour food frequency survey, asthma status, lung function test results, or other key covariates. The participant selection flow is illustrated in Figure 1, detailing exclusions due to missing data. This study was a retrospective analysis of existing KNHANES data. The researchers accessed the database for research purposes in July 2024. During and after the data collection process, the authors had no access to any information that could identify individual participants, ensuring complete anonymity.

**Fig 1.**
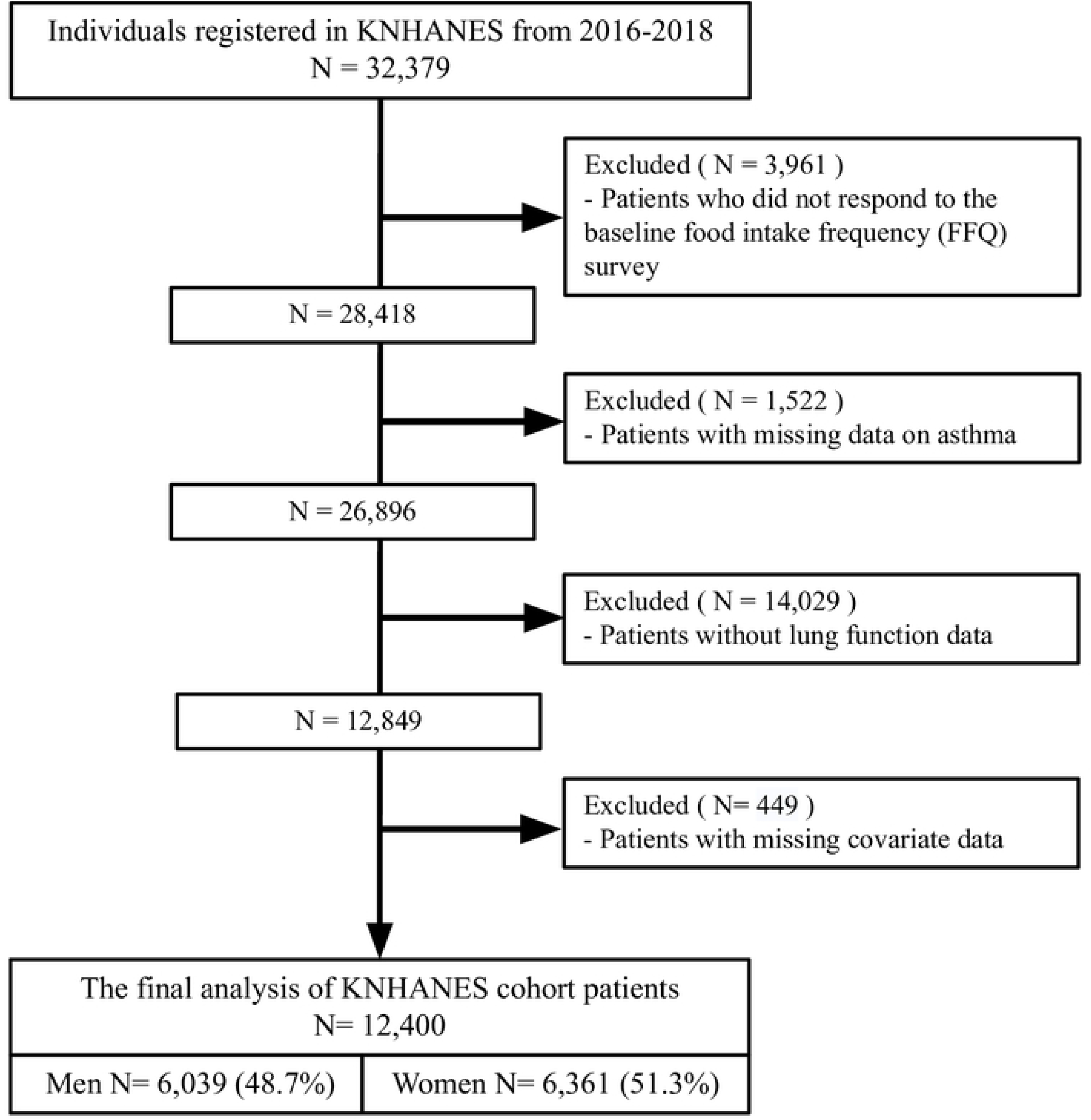
Flowchart of participant selection from the Korea National Health and Nutrition Examination Survey (KNHANES) 2016–2018. Among 32,379 individuals initially registered, exclusions were made for missing data on dietary intake, asthma status, lung function, and covariates. The final analysis included 12,400 participants (5,199 men and 7,228 women).

### Definition of Asthma and Measurement of Lung Function

Asthma status was assessed through self-reported responses to the following survey questions: (1) “Have you ever been diagnosed with asthma by a medical professional?” and (2) “Are you currently experiencing symptoms of asthma?”. Participants who answered “yes” to both questions were categorized into the current asthma group, while those who answered “no” to either were categorized into the non-asthma group. The operational definition of asthma used in KNHANES has been previously described [12]. Information on ever asthma was also collected based on self-reported physician diagnosis, regardless of current symptoms, to allow comparison with other definitions used in the literature.

Pulmonary function tests were conducted using a dry rolling seal spirometer (Vmax 2130, SensorMedics) in accordance with the American Thoracic Society/European Respiratory Society (ATS/ERS) guidelines [13]. The measurements reflect pre-bronchodilator values and were performed by trained technicians following standardized protocols during the Korea National Health and Nutrition Examination Survey (KNHANES) health examination (Korea Centers for Disease Control and Prevention, 2014) [14]. Lung function was assessed via forced expiratory volume in one second (FEV₁), forced vital capacity (FVC), the FEV₁/FVC ratio (%), and predicted values for FEV₁ and FVC.

### Assessment of Sociodemographic and Anthropometric Variables

Education level was categorized as elementary, middle school, high school, or college and above. BMI was calculated as weight (kg) divided by height squared (m²), and residence was classified as urban or rural.

### Assessment of Dietary Inflammatory Index (E-DII) and Nutrient Intake

Dietary intake was assessed through a single 24-hour dietary recall (24HR) conducted by trained dietitians during the KNHANES health interview. Participants were asked to recall all foods and beverages consumed during the 24 hours prior to the interview, typically from waking to bedtime. Nutrient intakes were calculated based on food items reported during the 24-hour dietary recall, using the Korean Food Composition Table, 9th revision [15, 16].

The DII was calculated according to the method developed by Shivappa et al. [8]. Z-scores for each dietary component were computed using the global standard mean and standard deviation, converted to centered percentile values, multiplied by corresponding inflammatory effect scores, and summed to obtain a total DII score. The E-DII was calculated by standardizing nutrient intake per 1000 kcal of total energy intake [9].

In this study, 21 food parameters were included: energy, carbohydrate, protein, total fat, saturated fat, monounsaturated fatty acids, polyunsaturated fatty acids, n-3 fatty acids, n-6 fatty acids, cholesterol, fiber, β-carotene, vitamin C, vitamin D, vitamin E, thiamin, riboflavin, niacin, magnesium, iron, and zinc.

### Statistical analysis

Descriptive statistics were used to summarize energy and nutrient intake, as well as E-DII scores, according to current asthma status. Differences in baseline characteristics between participants with and without asthma were assessed using the chi-squared test for categorical variables and the independent t-test for continuous variables.

To evaluate the relationship between E-DII scores and asthma, multiple logistic regression analyses were performed. Participants were also categorized into quartiles based on E-DII scores to assess trends in asthma prevalence across dietary inflammation levels. In the final models presented (Model 2), all regressions were adjusted for age and sex (male/female). In the descriptive presentation, sex was categorized as male or female, consistent with the KNHANES dataset. Additional sociodemographic variables were initially explored in preliminary models but were excluded from the final model to avoid overadjustment and multicollinearity.

In addition, linear regression analyses were conducted to examine the association between E-DII scores and lung function parameters, including FEV₁, FVC, and FEV₁/FVC ratio. All statistical analyses were performed using SAS software (version 9.4; SAS Institute, Cary, NC, USA) and were two-tailed. A *p*-value < 0.05 was considered statistically significant. For clarity, p-values < 0.001 were separately noted to emphasize strong associations.

## RESULTS

### Participant Characteristics by Asthma Status

**Table 1** summarizes the general characteristics of participants according to asthma status. Among 12,400 Korean adults, the mean age was 49.8 years and 51.3% were women. Of these, 246 (2.0%) reported current asthma and 12,154 (98.0%) did not. The asthma group had a significantly higher proportion of females (67.5% vs. 58.0%, *p* < 0.05) and older participants, particularly those aged ≥80 years (44.7% vs. 20.2%, *p* < 0.001). There were no significant differences in residence (urban vs. rural) or marital status between the two groups.

**Table 1.** Comparison of Sociodemographic and Health-Related Characteristics Between Asthma and Non-Asthma Groups.

|  |  | Asthma<br>(N=246) |  | Non-Asthma<br>(N=12,154) |  | *p*-value |
| --- | --- | --- | --- | --- | --- | --- |
| Sex | Male | 80 | 32.5 | 5110 | 42.0 | <0.05* |
|  | Female | 166 | 67.5 | 7044 | 58.0 |  |
| **Age** | 40-49 yr | 37 | 15.0 | 3261 | 26.8 | < 0.001** |
|  | 50-59 yr | 40 | 16.3 | 3372 | 27.7 |  |
|  | 60-69 yr | 59 | 24.0 | 3072 | 25.3 |  |
|  | ≥ 70 yr | 110 | 44.7 | 2449 | 20.2 |  |
| **Residence** | Urban | 107 | 43.5 | 5618 | 46.2 | 0.396 |
|  | Rural | 139 | 56.5 | 6536 | 53.8 |  |
| **Marital status** | Married | 243 | 98.8 | 11765 | 96.8 | 0.070 |
|  | Single | 3 | 1.2 | 389 | 3.2 |  |
| **Income** | Q1 | 94 | 38.2 | 2502 | 20.6 | < 0.001** |
|  | Q2 | 60 | 24.4 | 3007 | 24.7 |  |
|  | Q3 | 40 | 16.3 | 3122 | 25.7 |  |
|  | Q4 | 52 | 21.1 | 3523 | 29.0 |  |
| **Education level** | Elementary or lower | 117 | 47.6 | 2942 | 24.2 | < 0.001** |
|  | Secondary | 33 | 13.4 | 1601 | 13.2 |  |
|  | High school | 47 | 19.1 | 3882 | 31.9 |  |
|  | Bachelor's degree or higher | 49 | 19.9 | 3729 | 30.7 |  |
| **BMI (kg/m<sup>2</sup>)**<br>(mean ± SD) |  | 24.8 | 3.4 | 24.1 | 3.2 | <0.05* |
| **Alcohol** | No alcohol consumption | 58 | 23.6 | 1559 | 12.8 | < 0.001** |
|  | Current alcohol consumption | 129 | 52.4 | 8391 | 69.0 |  |
|  | Former alcohol consumption | 59 | 24.0 | 2204 | 18.1 |  |
| **Smoking** | Never smoker | 153 | 62.2 | 7383 | 60.8 | 0.248 |
|  | Current smoker | 28 | 11.4 | 1833 | 15.1 |  |
|  | Ex-smoker | 65 | 26.4 | 2938 | 24.2 |  |

\* $p < 0.05$ ; \*\* $p < 0.001$

Income level and education showed significant differences. The asthma group had a higher proportion of individuals in the lowest income quartile (38.2% vs. 20.6%) and a lower proportion in the highest quartile (21.1% vs. 29.0%) (*p* < 0.001). Educational attainment was also lower among asthma participants, with a greater proportion having elementary education or less (47.6% vs. 24.2%) and fewer having a bachelor’s degree or higher (19.9% vs. 30.7%) (*p* < 0.001).

Mean BMI was significantly higher in the asthma group (24.81 ± 3.43 kg/m²) than in the non-asthma group (24.17 ± 3.25 kg/m²) (*p* < 0.05). Alcohol consumption patterns differed significantly, with more no alcohol consumption and fewer current alcohol consumption among participants with asthma.

In addition, **Supplementary Table 1** presents a comparison of dietary intake between asthma and non-asthma groups. Participants with asthma had a more unfavorable nutrient profile, characterized by lower intake of dietary fiber, polyunsaturated fatty acids (PUFA), n-3 fatty acids, and vitamin C, while showing higher intake of total fat, saturated fat, and cholesterol.

### Association between E-DII and Current Asthma

**Table 2** presents the association between the E-DII and current asthma, analyzed using two models. Model 1 is a crude model without adjustments, while Model 2 is adjusted for sex and age. When treated as a continuous variable, E-DII showed a significant positive association with current asthma in both models (Model 1: odds ratio [OR] = 1.093, 95% CI: 1.048–1.139, *p* < 0.001; Model 2: OR = 1.079, 95% CI: 1.033–1.127, *p* < 0.001). When categorized into quartiles, the reference group was Quartile 1 (–6.600 to –3.689). In Model 1, compared to Quartile 1, the odds ratios for Quartiles 2, 3, and 4 were 1.158 (95% CI: 0.775–1.732), 1.363 (95% CI: 0.924–2.009), and 1.983 (95% CI: 1.388–2.851), respectively. A statistically significant association was observed only for Quartile 4 (*p* < 0.001). In Model 2, the corresponding odds ratios were 1.108 (95% CI: 0.739–1.661), 1.263 (95% CI: 0.850–1.876), and 1.778 (95% CI: 1.218–2.596), with significance retained only for Quartile 4 (*p* < 0.05). Additionally, a significant linear trend was observed across quartiles in both models (Model 1: *p*-trend < 0.001; Model 2: *p*-trend < 0.05), suggesting a dose–response relationship between higher E-DII and asthma risk.

**Table 2.** Association Between Energy-Adjusted Dietary Inflammatory Index (E-DII) and Current Asthma: Crude and Adjusted Models.

|  | Model 1 |  |  |  | Model 2 |  |  |  |
| --- | --- | --- | --- | --- | --- | --- | --- | --- |
|  | OR | 95% CI | p-value | p-trend | OR | 95% CI | p-value | p-trend |
| E-DII (continuous) | 1.093 | 1.048 – 1.139 | < 0.001** | < 0.001** | 1.079 | 1.033 – 1.127 | < 0.001** | < 0.001** |
| E-DII (quartile) |  |  |  |  |  |  |  |  |
| Quartile 1<br>(-6.600 – -3.689) | 1 (ref.) |  |  |  | 1 (ref.) |  |  |  |
| Quartile 2<br>(-3.688 – -1.526) | 1.158 | 0.775 – 1.732 | 0.474 |  | 1.108 | 0.739 – 1.661 | 0.620 |  |
| Quartile 3<br>(-1.525 – 1.429) | 1.363 | 0.924 – 2.009 | 0.118 | < 0.001** | 1.263 | 0.850 – 1.876 | 0.248 | <0.05* |
| Quartile 4<br>(1.430 – 5.144) | 1.983 | 1.380 – 2.851 | < 0.001** |  | 1.778 | 1.218 – 2.596 | <0.05* |  |

\* $p < 0.05$ ; \*\* $p < 0.001$

In addition, participants with asthma had significantly higher energy-adjusted E-DII scores compared to those without asthma (0.07 ± 0.15 vs. 0.03 ± 0.16, *p* < 0.001), indicating a more pro-inflammatory dietary pattern (**Supplementary Table 2**). Consistently, across most nutrients, the asthma group showed lower intake of anti-inflammatory components (e.g., fiber, PUFA, n-3 fatty acids, and vitamin C) and higher intake of E-DII–contributing foods.

### Association between E-DII and Lung Function Parameters

Linear regression analysis was performed to assess the relationship between the E-DII and various lung function parameters, as shown in **Table 3**.

**Table 3.**
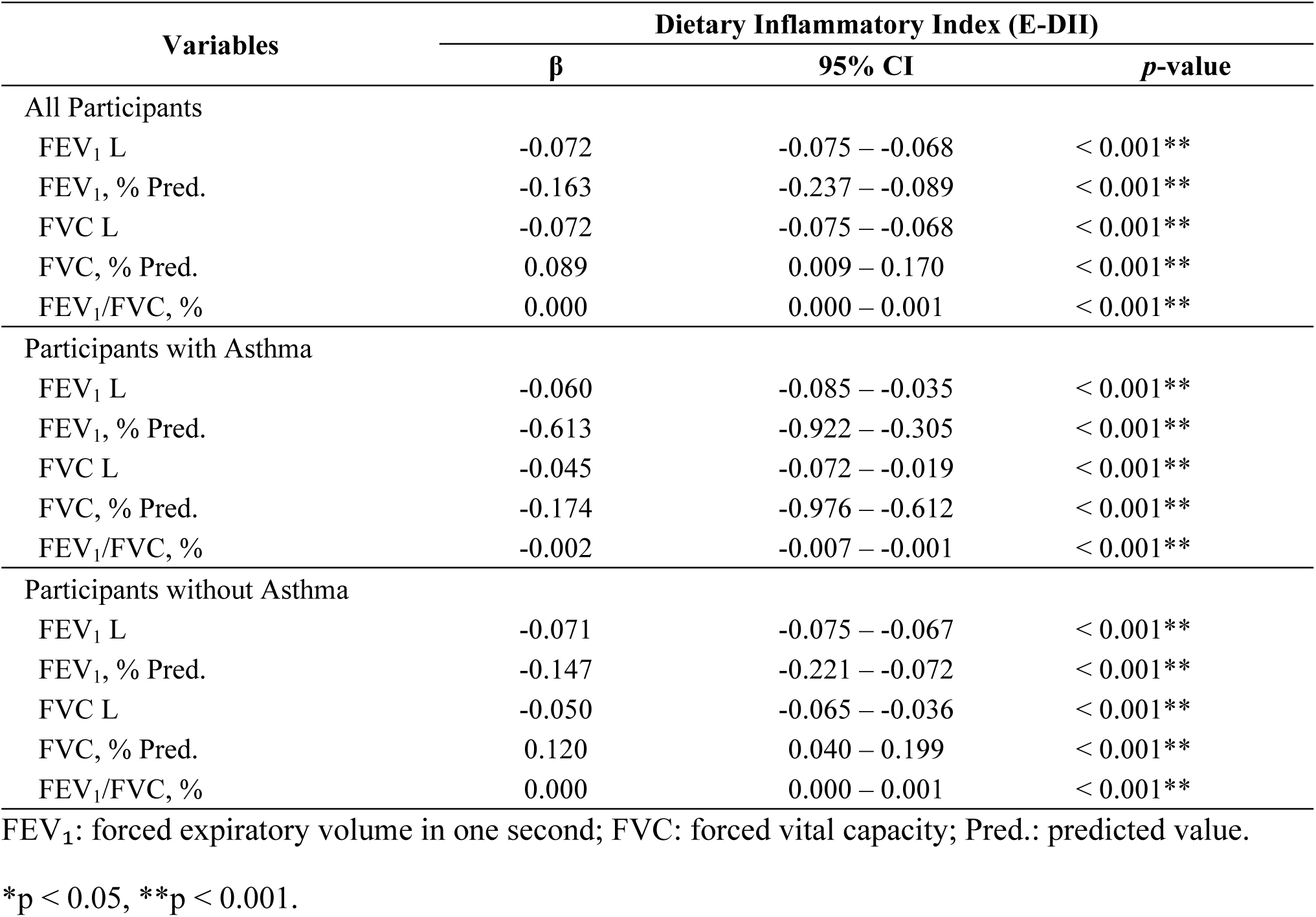
Association Between Energy-Adjusted Dietary Inflammatory Index (E-DII) and Lung Function Parameters in All Participants, and in Subgroups With and Without Asthma.

Among all participants, higher E-DII scores were significantly associated with lower FEV_1_ (β = −0.072, 95% CI: −0.075 to −0.068, *p* < 0.001), lower FEV_1_ % predicted (β = −0.163, 95% CI: −0.237 to −0.089, *p* < 0.001), and lower FVC (β = −0.072, 95% CI: −0.075 to −0.068, *p* < 0.001). FVC % predicted showed a significant positive association (β = 0.089, 95% CI: 0.009 to 0.170, *p* < 0.001), while FEV_1_/FVC ratio also showed a minimal but statistically significant positive association (β = 0.000, 95% CI: 0.000 to 0.001, *p* < 0.001).

In participants with asthma, higher E-DII scores were significantly associated with lower FEV1 (β = −0.060, 95% CI: −0.085 to −0.035, p < 0.001) and lower FVC (β = - 0.045, 95% CI: −0.072 to −0.019, p < 0.001). In participants with asthma, higher E-DII scores were significantly associated with a substantial decrease in $FEV_1$ % predicted ($\beta$ = −0.613, 95% CI: −0.922 to −0.305, $p < 0.001$) and lower FVC % predicted.

Among participants without asthma, the associations remained significant and consistent: FEV_1_ (β = −0.071, 95% CI: −0.075 to −0.067, *p* < 0.001), FEV_1_ % predicted (β = −0.147, 95% CI: −0.221 to −0.072, *p* < 0.001), and FVC (β = −0.050, 95% CI: −0.065 to −0.036, *p* < 0.001) all declined with increasing E-DII. FVC % predicted increased with E-DII (β = 0.120, 95% CI: 0.040 to 0.199, *p* < 0.001), and FEV_1_/FVC ratio also showed a small but significant positive association.

Logistic regression analysis was used to examine the association between the energy-adjusted Dietary Inflammatory Index (E-DII) and the likelihood of having asthma, as shown in **Table 4**.

**Table 4.**
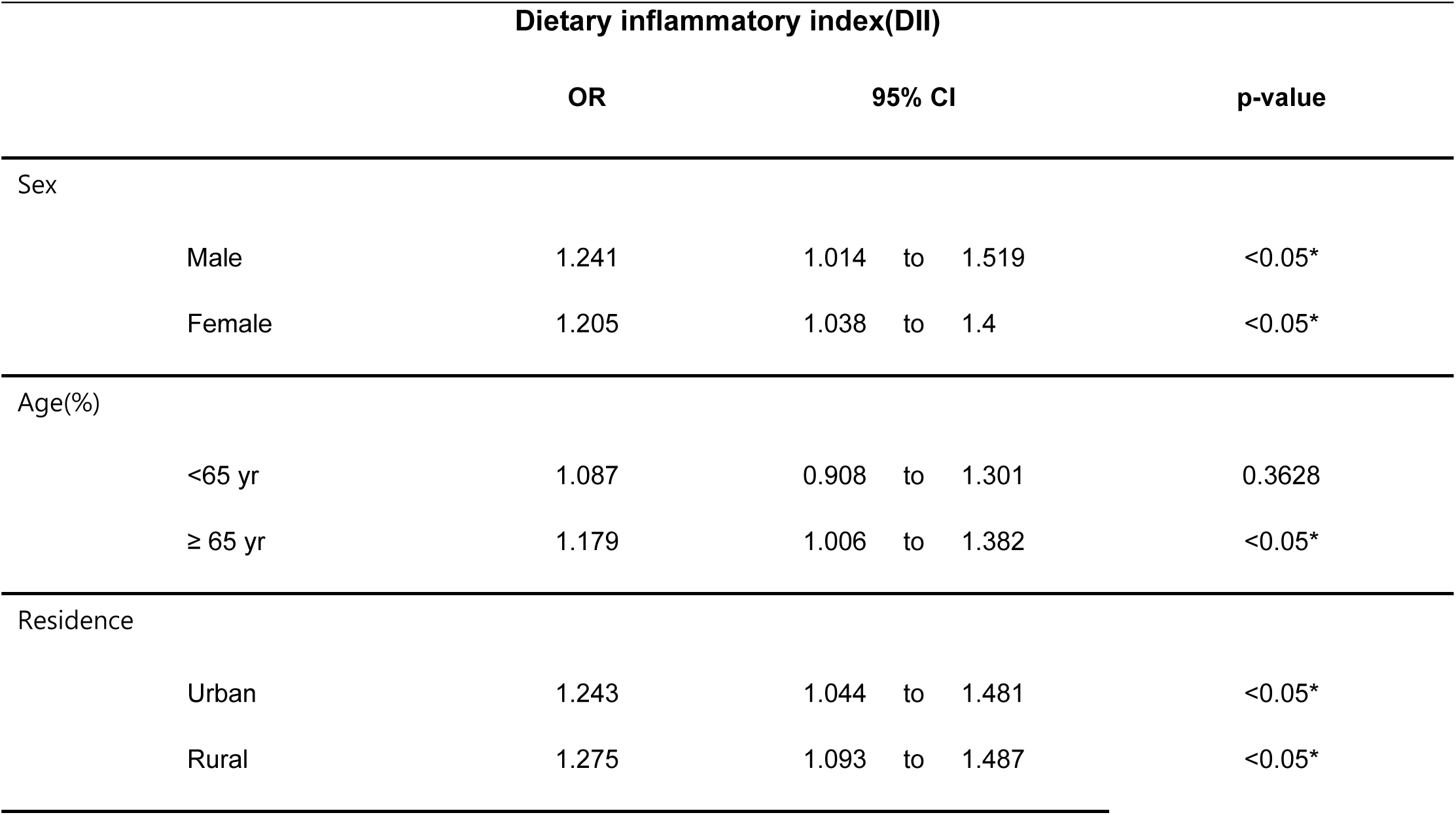
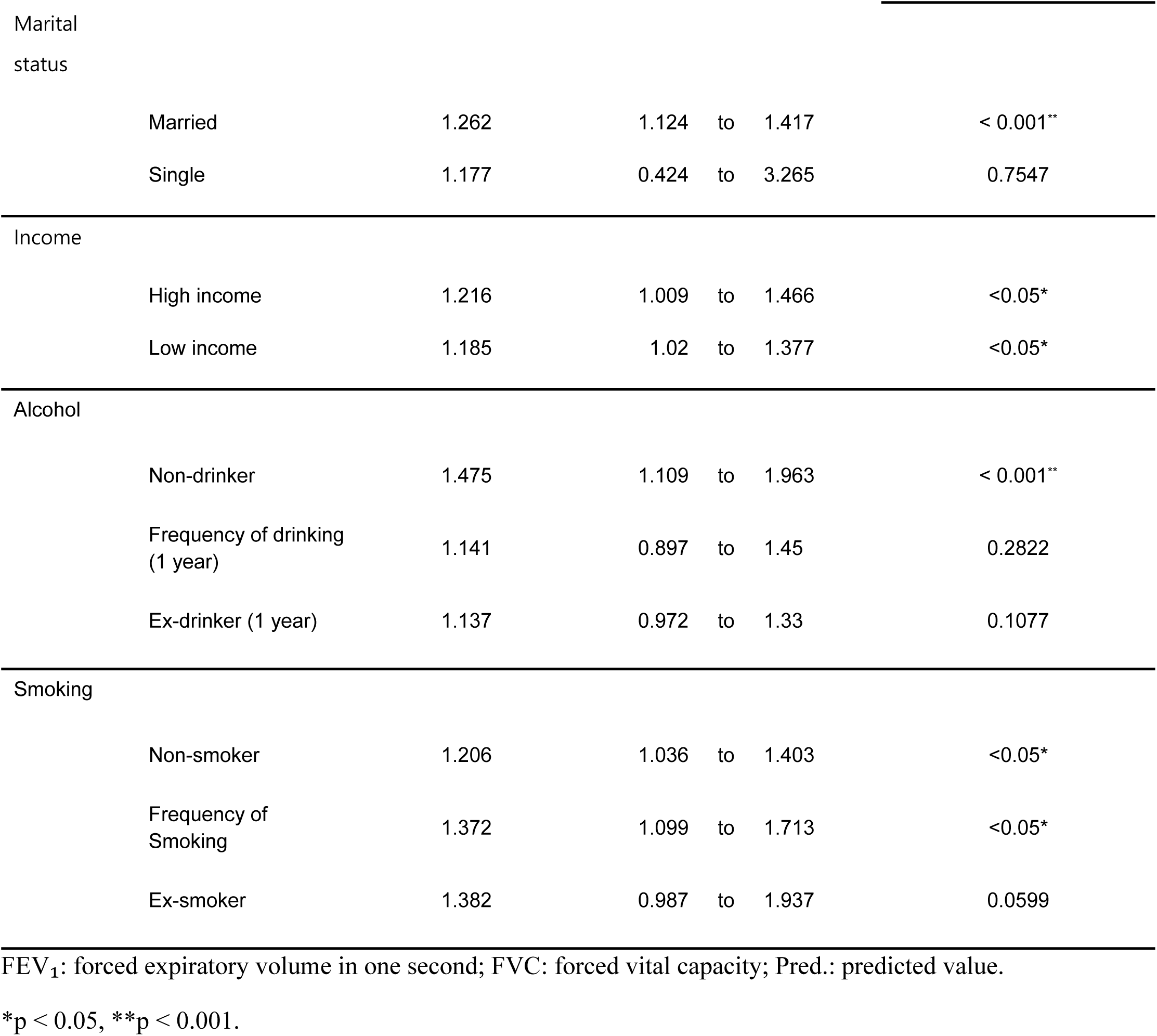
The energy-adjusted Dietary Inflammatory Index (E-DII) by relevant covariates in current asthma.

Higher E-DII scores were significantly associated with increased odds of asthma in several subgroups. For rural residents, a significant trend was observed, with higher E-DII scores showing increased odds of asthma (OR = 1.275, 95% CI: 1.093 to 1.487, p < 0.05). Similarly, married individuals exhibited a significant trend, with higher E-DII scores associated with increased odds of asthma (OR = 1.262, 95% CI: 1.124 to 1.417, p < 0.001). Among low-income participants, higher E-DII scores were associated with increased odds of asthma (OR = 1.185, 95% CI: 1.020 to 1.377, p < 0.05). Non-drinkers also demonstrated a significant trend, with higher E-DII scores associated with increased odds of asthma (OR = 1.475, 95% CI: 1.109 to 1.963, p < 0.001). Non-smokers and ex-smokers showed similar patterns, with higher E-DII scores linked to increased odds of asthma. For non-smokers, the trend was significant (OR = 1.206, 95% CI: 1.036 to 1.403, p < 0.05), and for ex-smokers, the odds were notably higher with increased E-DII scores (OR = 1.382, 95% CI: 0.987 to 1.937, p = 0.059).

## DISCUSSION

This study demonstrated a positive association between the E-DII score and the prevalence of asthma, with statistically robust associations consistent across models. As shown in baseline characteristics (**Table 1**), individuals with asthma tended to be female, older, of lower income and education level, with higher BMI, and were less likely to be current alcohol consumers compared with the non-asthma group. These findings suggest that dietary inflammation may be associated with asthma risk in a population-specific context and align with a recent NHANES-based study that reported higher DII scores to be associated with increased asthma prevalence in US adults [17].

Higher E-DII scores were also associated with lower FEV₁ and FVC levels. Although the absolute effect sizes were modest, the consistent direction and statistical robustness suggest a potential cumulative impact of dietary inflammation on pulmonary function at the population level. For instance, the β coefficient of –0.072 L for FEV₁ corresponds to roughly 2–3% of the average FEV₁ in Korean adults (2.5–3.5 L). While unlikely to be clinically significant for an individual, such changes could increase the proportion of the population falling below clinically important thresholds for impaired lung function.

Participants with asthma demonstrated significantly lower intake of total energy, macronutrients (carbohydrates, fat, and protein), and several micronutrients (e.g., β-carotene, vitamins C, D, and E, magnesium, and zinc) compared with those without asthma. These reductions may reflect disease-related changes in dietary habits, including avoidance of symptom-triggering foods, reduced appetite, or medication effects. Such alterations in nutrient intake are consistent with previous studies reporting modified dietary patterns among individuals with asthma [18] and may partly explain the higher inflammatory potential of their diets, as indicated by significantly higher E-DII scores in the asthma group. Notably, the magnitude of the association between E-DII and $FEV_1$ % predicted was markedly stronger in the asthma group (β = –0.613) than in the non-asthma group (β = –0.147). This disparity suggests that individuals with pre-existing airway inflammation may be more susceptible to the adverse effects of a pro-inflammatory diet, potentially leading to more pronounced lung function impairment compared to the general population. Mechanistically, specific dietary components can modulate immune responses relevant to asthma pathophysiology. Pro-inflammatory nutrients (e.g., saturated fats, cholesterol) may promote Th2-driven cytokine secretion (IL-4, IL-5, IL-13), aggravating allergic inflammation, whereas anti-inflammatory nutrients (e.g., unsaturated fats, vitamins, fiber) may promote Th1 and regulatory T cell responses, potentially mitigating allergic risk [6]. A previous meta-analysis demonstrated modest protective effects of vitamins C, E, and D against asthma and wheezing [5]. However, single-nutrient interventions such as vitamin D supplementation have shown limited preventive effects in randomized trials [19, 20]. These findings support the evaluation of overall dietary patterns, as captured by indices like the E-DII, which simultaneously account for anti- and pro-inflammatory dietary factors. This perspective is consistent with previous studies reporting protective effects of anti-inflammatory dietary patterns (e.g., Mediterranean diet) and harmful effects of Westernized diets on asthma and lung function [21–24]. Together with prior research [25–27], our results suggest that high dietary inflammation may be linked to increased asthma risk and reduced lung function, even when effect sizes are small.

The prevalence of current asthma in our dataset was 2.0%, consistent with national KNHANES estimates (2–3%) but lower than in Western countries (15–20%) [2]. This difference may reflect genetic factors or the anti-inflammatory nature of traditional Korean diets rich in vegetables and low in fat [28]. Our prevalence estimate was also slightly lower than the 3.15% reported by Kim et al. (2025) using the KNHANES 2018–2021 dataset [29]. This discrepancy likely reflects methodological differences, as our definition of “current asthma” required both a physician diagnosis and the presence of current symptoms at the time of survey, and we excluded participants with missing dietary intake, lung function, or key covariate data. These stricter criteria and exclusions should be considered when interpreting the prevalence estimates.

Some findings were counterintuitive. In non-asthma participants, E-DII showed positive associations with FVC% and the FEV₁/FVC ratio, which could reflect measurement variability, compensatory physiological mechanisms in those without airway disease, or heterogeneity within the non-asthma group. However, although these associations reached statistical significance, their clinical relevance is likely limited and may instead reflect subgroup characteristics, such as healthier individuals with higher dietary intake but without airway pathology. Similarly, although asthma participants showed a slightly lower proportion of current smokers than non-asthma participants, the difference was not statistically significant. This may be due to post-diagnosis smoking cessation, which classifies many as former rather than current smokers, the lack of lifetime smoking data, and possible response bias. These factors could have led to an underestimation of smoking prevalence among asthma patients.

This study has several limitations. First, as with other recall-based dietary assessments, a single 24-hour recall (24HR) is subject to recall bias and may not adequately represent an individual’s usual or long-term dietary patterns, as it captures only one day or a specific season of intake. This limitation can introduce measurement error or misclassification in estimating the dietary inflammatory index, potentially attenuating observed associations. Such misclassification is generally non-differential, biasing results toward the null; however, differential misclassification is also possible if recall accuracy varies by disease status. Validation studies in Korean populations have shown that a single 24HR exhibits a moderate level of correlation with long-term dietary assessment methods (e.g., repeated recalls or food frequency questionnaires) [30]. This suggests that a single 24HR may provide a reasonable approximation of overall dietary patterns, though not a complete one. Therefore, future research incorporating repeated recalls or validated food frequency questionnaires is warranted to obtain more accurate estimates.

Second, not all nutrients in the original E-DII were available, and IL-6 data was missing, potentially affecting score precision. Third, potential confounders such as medication use and environmental exposures were not controlled. Due to the lack of detailed medication data in KNHANES, we could not assess associations between E-DII and asthma treatments such as corticosteroids. Moreover, despite efforts to select covariates with clinical and epidemiological relevance, only age and sex were included in the final models, and residual confounding by unmeasured or inadequately measured factors cannot be excluded. Future studies with linked clinical treatment records and more comprehensive covariate adjustment are needed to address these issues.

Fourth, asthma diagnosis was based on self-report, which may have led to misclassification despite the use of dual-definition criteria intended to improve specificity. This approach likely enhanced diagnostic accuracy, but some misclassification remains possible. For instance, individuals with a past asthma diagnosis but no current symptoms may have been excluded, while symptomatic individuals without a formal diagnosis, particularly those with limited healthcare access, may have been underrepresented. In addition, reported symptoms could reflect other respiratory or cardiac conditions. Finally, the absence of objective diagnostic tools such as spirometry or methacholine challenge testing in KNHANES limited diagnostic precision, and sensitivity analyses using alternative definitions of asthma could not be conducted.

Fifth, all regression models were adjusted for age and sex; however, additional stratified analyses or interaction tests by sex, age, or other characteristics could not be performed due to data limitations. This may restrict the interpretability and generalizability of our findings. In particular, although there was a notable difference in the proportion of participants aged ≥ 70 years between the asthma and non-asthma groups, a sensitivity analysis excluding this age group could not be conducted, which represents another limitation. Finally, selection bias is possible, as participants with incomplete data were excluded. Baseline covariate information for excluded individuals was unavailable, preventing direct comparison with the sample included

Despite these limitations, this is the first large-scale study in an Asian population to assess the association between E-DII, asthma, and lung function. While the cross-sectional design constrains causal inference, the study contributes to hypothesis generation in a population where such evidence has been scarce. By demonstrating consistent associations in this cohort, our findings complement previous reports largely derived from Western populations and emphasize the need for longitudinal and mechanistic studies. These findings highlight the potential role of dietary inflammation in asthma pathophysiology and suggest that dietary interventions could be considered in preventive strategies.

## CONCLUSIONS

In this nationally representative Korean adult population, higher E-DII scores were significantly associated with increased odds of current asthma and reduced lung function, after adjustment for age and sex. These findings suggest that pro-inflammatory dietary patterns may be associated with asthma pathogenesis and pulmonary impairment. While the observed effect sizes were modest, the associations were consistent and support the growing body of evidence linking diet and respiratory health. Given the modifiable nature of dietary habits, our results underscore the potential utility of dietary intervention strategies in asthma prevention and respiratory function maintenance.

Further longitudinal and interventional studies are warranted to establish causality and to explore the biological mechanisms underlying these associations.

## Data Availability

The data underlying the results presented in the study are available from the Korea National Health and Nutrition Examination Survey (KNHANES) conducted by the Korea Disease Control and Prevention Agency (KDCA). The datasets are publicly available and can be accessed through the official KNHANES website (https://knhanes.kdca.go.kr/knhanes/eng/index.do) upon request and approval for research purposes.

## Acknowledgments

This research was supported by the Gangwon RISE Center, funded by the Ministry of Education and Gangwon State (2025-RISE-10-009).

## Supporting Information Captions

S1 Table. Comparison of food intake between Asthma Group and Non-Asthma group.

